# Convergence of HIV and non-communicable disease epidemics: Geospatial mapping of the unmet health needs in a HIV Hyperendemic South African community

**DOI:** 10.1101/2023.03.27.23287807

**Authors:** Diego F Cuadros, Chayanika Devi, Urisha Singh, Stephen Olivier, Alison Castle, Yumna Moosa, Johnathan A Edwards, Hae-Young Kim, Mark J. Siedner, Emily B Wong, Frank Tanser

**Author notes:** Reprints or correspondence: Diego F. Cuadros, Ph.D., Digital Epidemiology Laboratory, Digital Futures, University of Cincinnati, Cincinnati, OH, 45221.

## Abstract

**Background:** As people living with HIV (PLHIV) are experiencing longer survival, the co-occurrence of HIV and non-communicable diseases has become a public health priority. In response to this emerging challenge, we aimed to characterize the spatial structure of convergence of chronic health conditions in a HIV hyperendemic community in KwaZulu-Natal, South Africa.

**Methods:** We utilized data from a comprehensive population-based disease survey conducted in KwaZulu-Natal, South Africa, which collected data on HIV, diabetes, and hypertension. We implemented a novel health needs scale to categorize participants as: diagnosed and well-controlled (Needs Score 1), diagnosed and sub-optimally controlled (Score 2), diagnosed but not engaged in care (Score 3), or undiagnosed and uncontrolled (Score 4). Scores 2-4 were indicative of unmet health needs. We explored the geospatial structure of unmet health needs using different spatial clustering methods.

**Findings:** The analytical sample comprised of 18,041 individuals. We observed a similar spatial structure for HIV among those with a combined needs Score 2-3 (diagnosed but uncontrolled) and Score 4 (undiagnosed and uncontrolled), with most PLHIV with unmet needs clustered in the southern peri-urban area, which was relatively densely populated within the surveillance area. Multivariate clustering analysis revealed a significant overlap of all three diseases in individuals with undiagnosed and uncontrolled diseases (unmet needs Score 4) in the southern part of the catchment area.

**Interpretation:** In a HIV hyperendemic community in South Africa, areas with the highest needs for PLHIV with undiagnosed and uncontrolled disease are also areas with the highest burden of unmet needs for other chronic health conditions, such as diabetes and hypertension. The identification and prioritization of geographically clustered vulnerable communities with unmet health needs for both HIV and non-communicable diseases provide a basis for policy and implementation strategies to target communities with the highest health needs.

**Funding:** Research reported in this publication was supported by the Fogarty International Center (R21 TW011687; D43 TW010543), the National Institute of Mental Health, the National Institute of Allergy and Infectious Diseases (K24 HL166024; T32 AI007433) of the National Institutes of Health, and Heart Lung and Blood Institute (K24 HL166024, T32 AI007433). The contents of this manuscript are solely the responsibility of the authors and do not necessarily represent the official views of the funders.

## INTRODUCTION

While the transition of disease burden has predominantly included shifts from infectious diseases to non-communicable diseases (NCDs) globally, numerous studies in South Africa (SA) have also reported the potential emergence of multimorbidity interactions with the convergence of infectious diseases and NCDs.^1-6^ Likewise, life expectancy in people living with HIV (PLHIV) has also rebounded following the increasing access to antiretroviral therapy (ART), resulting in an increased likelihood of comorbidities in this vulnerable population.^7,8^ As a result, detailed epidemiologic studies of colliding HIV and NCDs in SA remain an important and urgent priority to improve the delivery of targeted healthcare.^9^

Recent applications of mapping and spatial analysis have proven to be an effective methodology to uncover health determinants and vulnerable populations at higher risk of chronic health conditions.^10,11^ However, several studies aimed to uncover the geospatial convergence of different diseases have found no evidence of geographical overlap between the prevalence of HIV and NCDs like cardiovascular diseases and diabetes in SA, at country and at locale level.^6,12,13^ These results were consistent with other studies that also found no epidemiological link between PLHIV and the presence of any NCDs in SA and in other countries in sub-Saharan Africa (SSA).^14-16^ Differential local biological, demographic, and socioeconomic determinants driving HIV and NCDs microepidemics could be preventing the detection of the overlap among these health chronic conditions in the same communities. Likewise, distinct epidemic stages might generate a mismatch in the epidemic intensities of the diseases that are colliding in the same communities, resulting in lack of awareness about multimorbidity.

Understanding disease progression and the unmet health needs of individuals with chronic conditions is an important step to addressing disease co-occurrence and designing health system response to these. The extent to which health needs of different diseases overlap within individuals and communities would help to uncover disease interactions emerging from multimorbidity associations. In the Vukuzazi study, we assessed the health needs for HIV, hypertension, and diabetes in a HIV hyperendemic community in Kwazulu-Natal, SA.^17^ In this study we introduced a novel health needs framework to conceptualize the unmet health needs of communities impacted by the overlapping infectious and noninfectious diseases in the region.

Applying this framework, this study found that half of the people living with HIV, hypertension, or diabetes in this community have unmet health needs, with the greatest concentration of unmet health needs among persons living with NCDs. Furthermore, preliminary spatial visualizations demonstrated unanticipated geographical overlap of unmet health needs that could be used to inform strategies of health service delivery.

In the present study, we build upon prior research by conducting in-depth geospatial analyses of the spatial distribution of unmet health needs for HIV, hypertension, and diabetes in KwaZulu-Natal, SA. Leveraging a health needs framework previously developed and employing various spatial epidemiology and disease mapping techniques, we examined the geographic convergence of unmet health needs and identified areas where individuals with the greatest needs for HIV and NCDs are concentrated. The identification of these spatial patterns holds important implications for designing prevention and monitoring strategies for individuals with the highest health needs, which could optimize interventions aimed at enhancing services for these vulnerable communities at higher risk of overlapping health conditions.

## METHODS

### Data sources

We analyzed data collected in the Vukuzazi Study that was conducted in the uMkhanyakude district of KwaZulu-Natal, SA between 2018-2020. Sample population and data collection have been described in detail elsewhere.^6^ Briefly, eligible adolescent and adult residents, >15 years of age, from the Africa Health Research Institute (AHRI) Demographic Surveillance Area (DSA) in the uMkhanyakude district of KwaZulu-Natal located near the market town of Mtubatuba and covers 438□km^2^, were invited to participate in this cross-sectional survey.^18^ Participants were visited at their homesteads and invited to participate at a mobile health camp that moved sequentially through the study area during the study period. Anthropomorphic and blood pressure measurements were performed using the World Health Organization STEPS protocol.^19^ Blood was collected for measurement of glycosylated hemoglobin (HbA1c, VARIANT II TURBO Haemoglobin testing system (Bio-Rad, Marnes-la-Coquette, France)) and for HIV (Genscreen Ultra HIV Ag-Ab enzyme immunoassay (Bio-Rad)). For this analysis, we used the health needs definitions proposed by Singh et al.,^17^ in which a health needs score for each disease was assigned as following: (i) diagnosed and optimally treated (Score 1), (ii) diagnosed and sub-optimally treated (Score 2), (iii) diagnosed but not engaged in care (Score 3), and (iv) undiagnosed but with a positive screening test in Vukuzazi (Score 4). Detailed health definitions for each score are included in Table 1. For this study, we focused our analyses in the unmet health needs (Score 2 to 4), and combined Score 2 and 3 for individuals diagnosed in need of treatment, and Score 4 for individuals with the highest needs. This study follows the Strengthening the Reporting of Observational Studies in Epidemiology (STROBE) reporting guideline.^20^

**Table 1.** Need score definitions for HIV, diabetes, and hypertension

| Need Score | HIV | Diabetes | Hypertension (HPTN) |
| --- | --- | --- | --- |
| **Score 1:** Diagnosed, engaged in care, and optimally treated | Known diagnosis of HIV<br>On treatment<br>HIV viral load < 40 | Known diagnosis of diabetes<br>On treatment<br>HBA1c ≤6.5 | Known diagnosis of HPTN<br>on treatment<br>Blood pressure ≤140/≤90mmHg |
| **Score 2:** Diagnosed, engaged in care, and sub-optimally treated | Known diagnosis of HIV<br>On treatment<br>HIV viral load > 40 | Known diagnosis of diabetes<br>On treatment<br>HBA1c >6.5 | Known diagnosis of HPTN<br>on treatment<br>Blood pressure ≥140/≥90mmHg |
| **Score 3:** Diagnosed, not engaged in care | Known diagnosis of HIV<br>Not on treatment<br>HIV viral load > 40 | Known diagnosis of diabetes<br>not on treatment<br>HBA1c >6.5 | Known diagnosis of HPTN<br>not on treatment<br>Blood pressure ≥140/≥90mmHg |
| **Score 4:** Previously undiagnosed, positive screening test | No previous diagnosis of HIV<br>Immunoassay positive<br>HIV viral load >40 | No previous diagnosis of diabetes<br>HBA1c ≥6.5 | No previous diagnosis of HPTN<br>Blood pressure ≥140/≥90mmHg |

### Geospatial analyses

Participants included in the study were geolocated to their respective homesteads of residence using the comprehensive geographic information system. To protect the confidentiality of the participants a geographical random error was introduced to the geographical coordinates of each homestead included in the study.^21^ The geospatial distribution of the prevalence of needs Score 2 and 3 combined (individuals diagnosed in need of treatment), and needs Score 4 (individuals with the highest needs) for each of three diseases included in the study were assessed by the generation of continuous surface maps using a standard Gaussian kernel interpolation method (with a search radius of 3 km), which has been used and validated in this population for mapping multiple HIV outcomes in the area of study,^21^ and the spatial structure of the prevalence of these unmet needs scores for each disease was identified using optimized hotspot analysis, which identifies areas with high (hotspots) or low (coldspots) prevalence of these unmet needs.^22,23^ We identified geospatial associations between a combination of the needs score of two diseases applying spatial bivariate and multivariate analysis using the geospatial GeoDa environment.^24^ First, spatial correlations between a combination of the needs score of two diseases were identified using bivariate local indicators of spatial association (LISA). The bivariate LISA statistics identified significant spatial clustering based on the degree of linear association between the prevalence of the needs score of one disease at a given location and the prevalence of the needs score of the other disease at neighbouring locations.^25^ Maps were generated illustrating the locations with statistically significant associations and the type of spatial association between the needs score of both diseases (i.e. high–high, low–low, low–high, and high-low). Second, we implemented a method previously used to identify multivariable spatial associations of health determinants using K-means clustering analysis^13,26-29^ to identify the spatial relationship between the distribution of the prevalence of the highest needs (needs Score 4) of all three diseases, HIV, diabetes, and hypertension. K-means is a partitioning clustering method in which the data are partitioned into *k* groups (i.e., fourth groups). In this clustering method, the *n* observations are grouped into *k* clusters such that the intra-cluster similarity is maximized (or dissimilarity minimized), and the between-cluster similarity minimized (or dissimilarity maximized). A further detailed description of these geospatial methods can be found elsewhere.^30,31^ Percentages for the prevalence of needs score 4 for each disease in every cluster identified were reported. Maps of the results were generated using ArcGIS Pro 3.1.0.^32^

### Ethical considerations

The Vukuzazi study was approved by the University of KwaZulu-Natal (UKZN) Biomedical Research Ethics Committee (BREC) and the institutional review board of Mass General Brigham. Written consent was obtained from all participants.

## RESULTS

A total of 18,041 individuals were enrolled in the Vukuzazi study, representing 50% of the 36,097 eligible residents of the DSA. We estimated that 54.9% of the participants had at least one of the three diseases included in the study. Of those individuals with at least one disease, 61.7% had HIV, 17.6% had diabetes, and 46.6% had hypertension. Maps in Figure 1 illustrate the spatial distribution of need Scores 2 and 3 combined, and need Score 4 for each disease.

**Figure 1.**
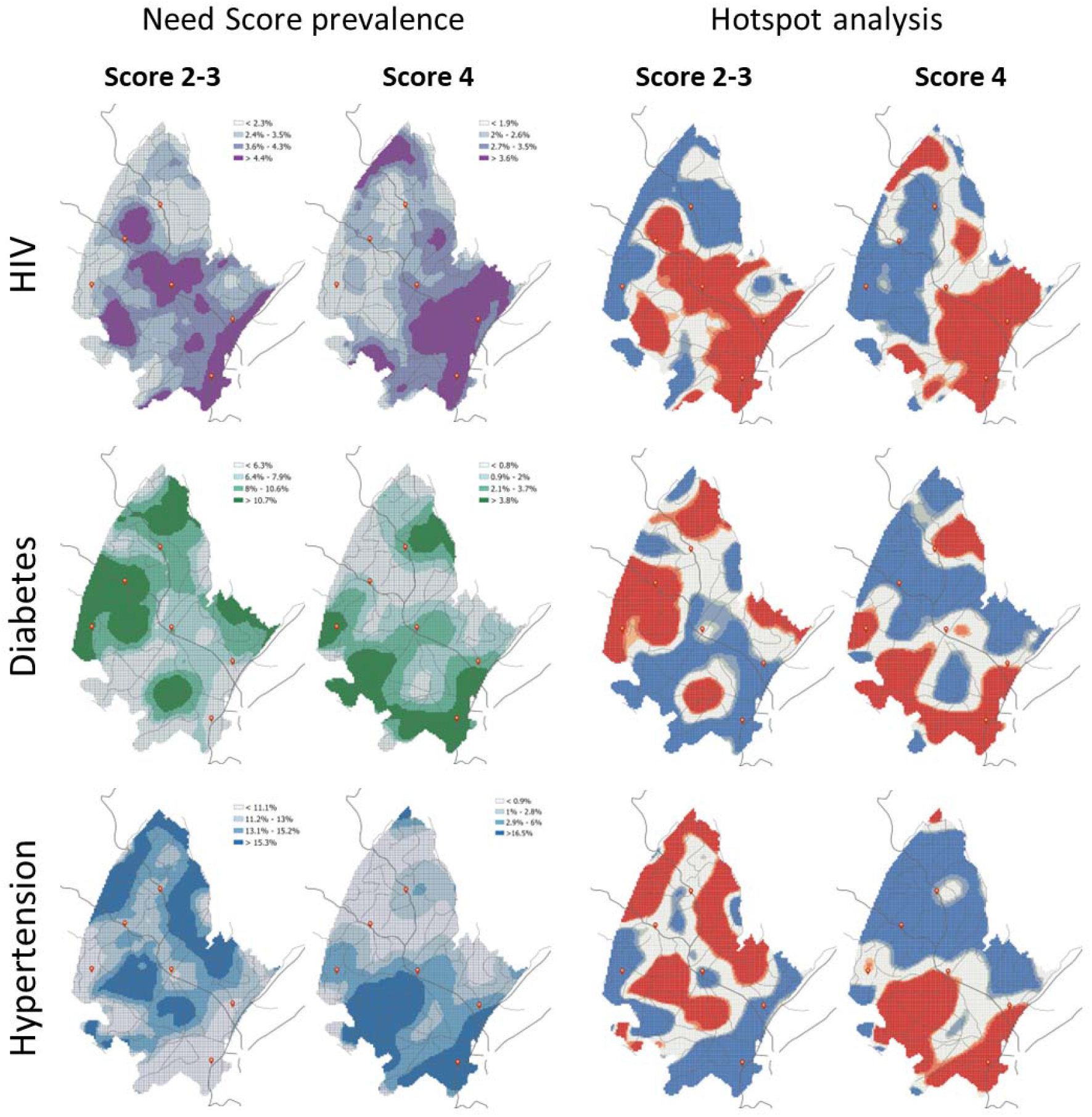
Spatial structure of the prevalence of need Scores 2 and 3 combine, and need Score 4 for each disease included in the study: HIV, diabetes, and hypertension. Maps on the right illustrate the results of the optimized hotspot analysis for each disease, with areas in red illustrating the identified hotspots, whereas areas in blue correspond to the location of the coldspots. Dark lines illustrate the main roads and red dots illustrate the location of the main healthcare facilities within the surveillance area

Need Scores 2 and 3, and need Score 4 had similar spatial distribution for HIV, with a higher concentration of individuals with these high needs concentrated in the southern part of the surveillance area. Conversely, a high prevalence of need Scores 2 and 3 for diabetes and hypertension were mostly distributed in the center and northern part of the surveillance area. Furthermore, a high prevalence of need Score 4 for diabetes and hypertension were mostly distributed in the southern part of the surveillance area. Estimated spatially smoothed prevalence for the different health need scores for all three diseases within the hotspots and coldspots identified are compared in Figure 2. Prevalence of need Score 4 for HIV within the hotspot was 3.9% compared to 1.5% within the identified coldspots. For diabetes, prevalence of need Score 4 within the hotspot was 5.0% compared to 0.8% within the coldspots, whereas prevalence of Score 4 for hypertension within the hotspot was 9.0% compared to 0.9% within the identified coldspots.

**Figure 2.**
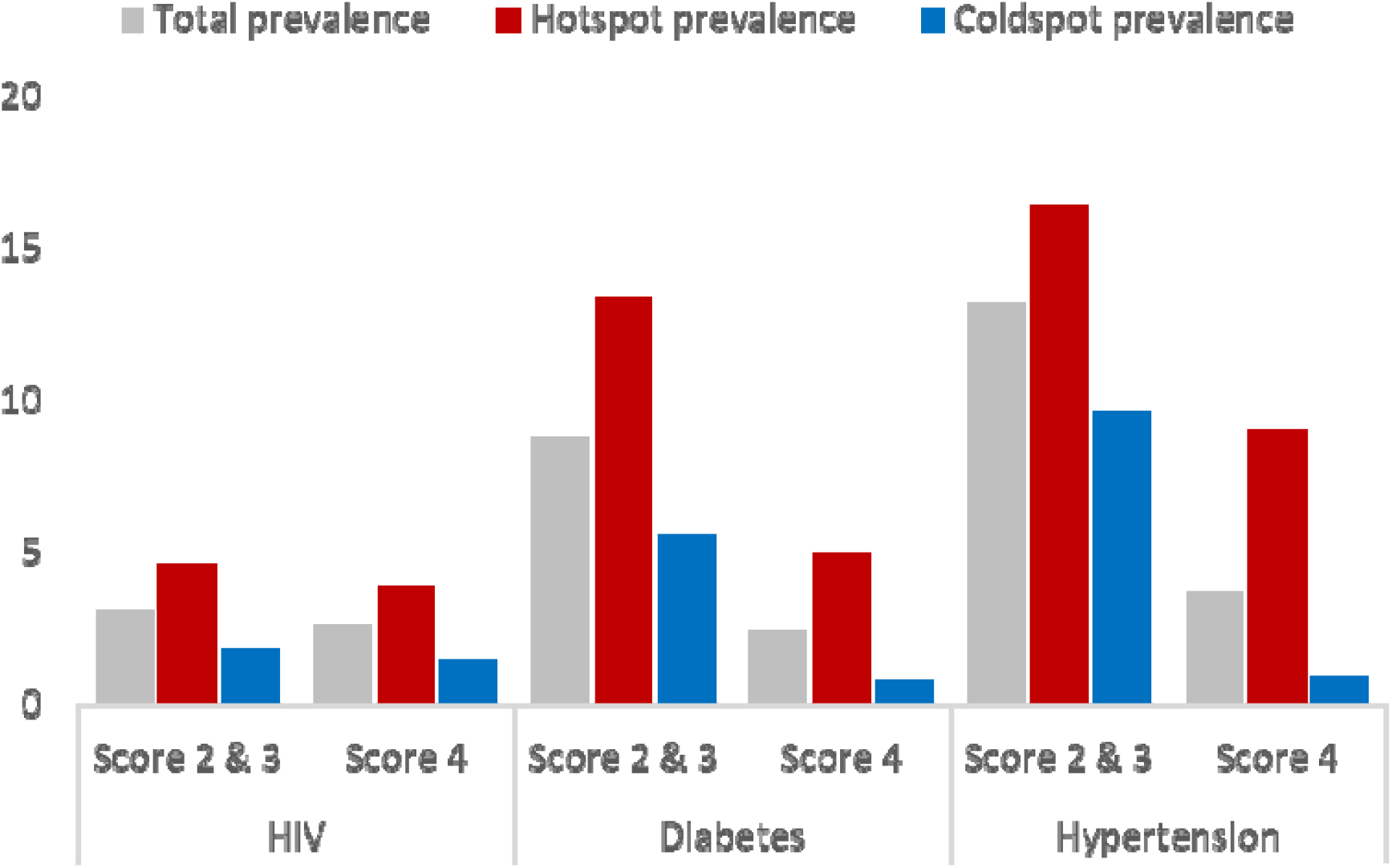
Estimated spatially smoothed prevalence for the need Scores 2 and 3 combine and, need Score 4 for all three diseases included in the study: HIV, diabetes, and hypertension. Grey bars illustrate the total prevalence for each disease, whereas red bars and blue bars illustrate the prevalence within the hotspot and the coldspot respectively

Bivariate LISA analysis did not identify a clear spatial pattern of associations between the combination need Scores 2 and 3 for all diseases (maps on top in Figure 3). Conversely, there was a significant association of overlapping Score 4 for all combination of diseases, which was located in the southern part of the surveillance area (green areas in maps of bottom of Figure 3). Multivariable K-means clustering analysis of need Score 4 was consistent with this result and identified a cluster (Cluster 4, dark blue areas in map in Figure 4A) located in the southern part of the surveillance area. This cluster had the highest prevalence of need Score 4 for diabetes (4.3%) and hypertension (8.2%), and the second highest prevalence of need Score 4 for HIV (3.2%). This cluster occupied 23.2% of the surveillance area and 52.1% (52,991 of the 101,627 individuals living in the DSA) ^18^ reside within this cluster with high prevalence of need Score 4 identified (Figure 4B).

**Figure 3.**
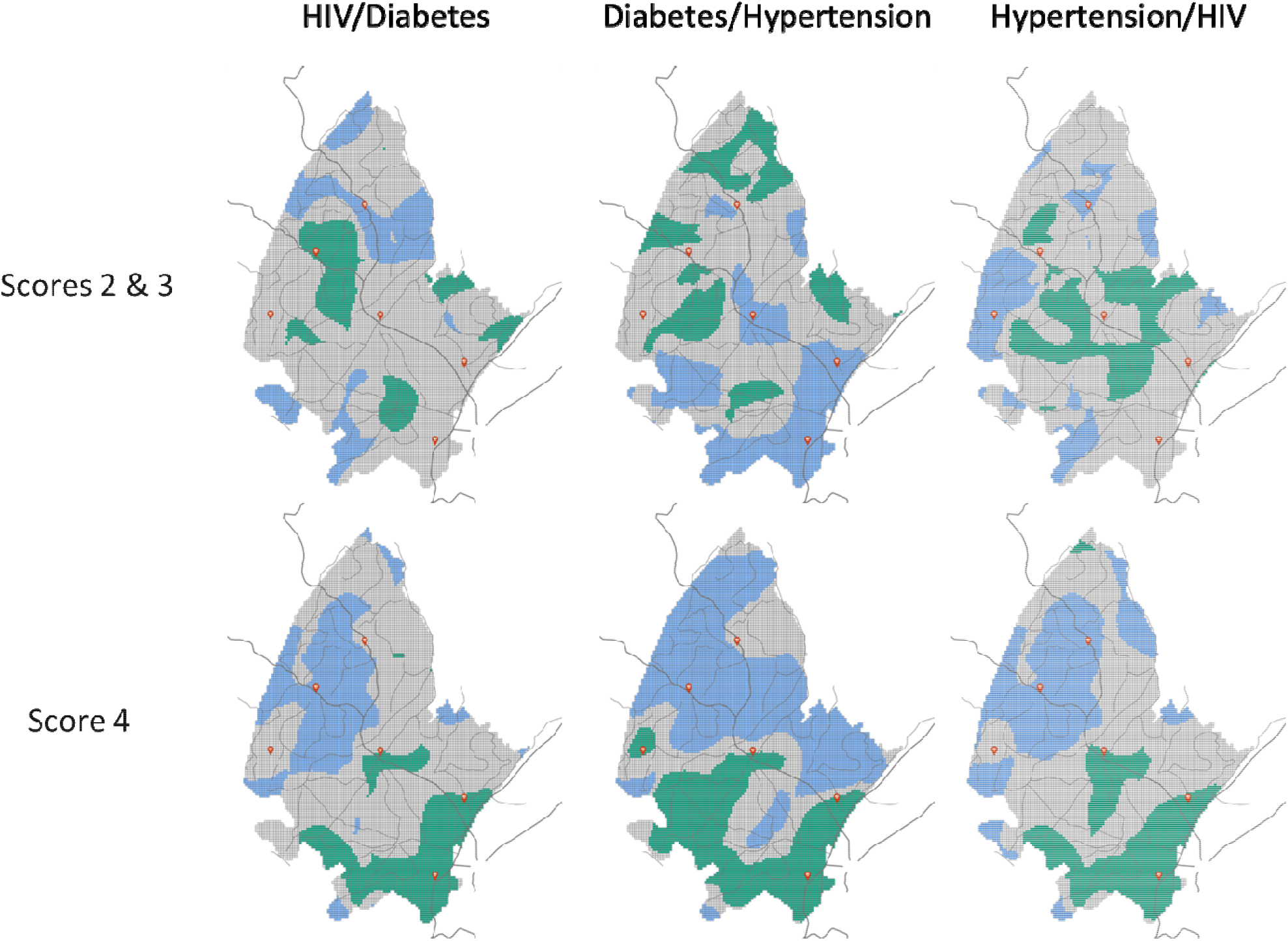
Linear association of spatial autocorrelation (LISA) analysis for the combination between the need scores for each disease included in the study. Green color indicates high-high association, whereas blue color indicates areas with low-low associations. Grey color indicated nonsignificant association. Dark lines illustrate the main roads and red dots illustrate the location of the main healthcare facilities within the surveillance area

**Figure 4.**
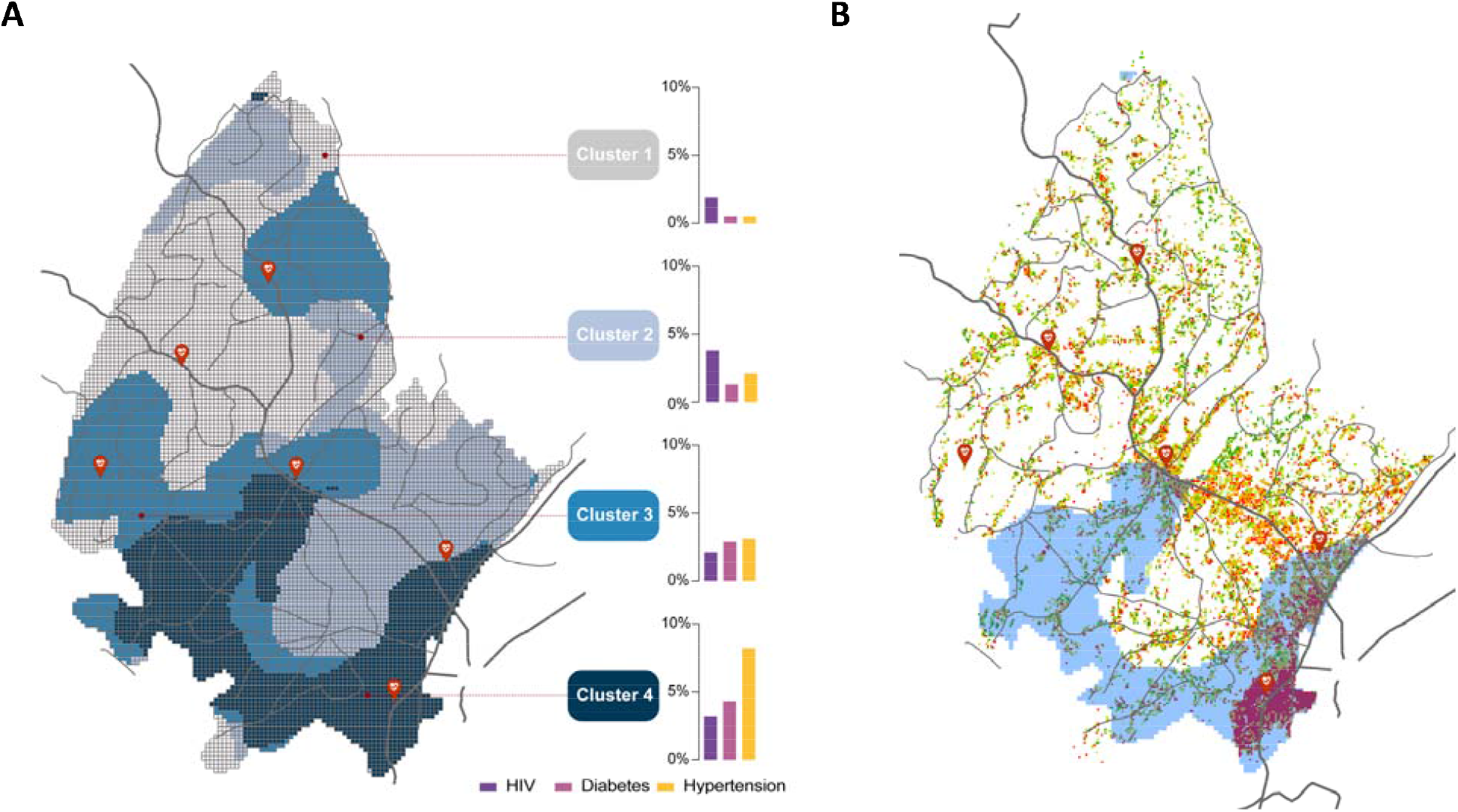
Multivariate K-means clustering analysis for need Score 4 of all three diseases combined (A). Cluster 4 with the highest prevalence of need Score 4 for diabetes and hypertension, and the second highest prevalence of need Score 4 for HIV is illustrated in dark blue. Population density distribution within the surveillance area and within Cluster 4 (B). Red areas illustrate highly dense population locations. Dark lines illustrate the main roads and red dots illustrate the location of the main healthcare facilities within the surveillance area

## DISCUSSION

In this study, we aimed to investigate the spatial distribution of health needs related to chronic health conditions, such as HIV, hypertension, and diabetes, in a community facing a hyperendemic HIV epidemic in SA. Our results showed that individuals with the highest health needs for these three diseases clustered in the same location within the surveillance area, indicating a significant overlap of unmet health needs. Multivariable clustering analysis confirmed this finding, revealing a cluster of individuals with the highest needs for all three diseases in the southern part of the surveillance area. This area, covering about 23% of the surveillance region, had the highest prevalence of undiagnosed and not engaged hypertension and diabetes care, and the second highest prevalence of PLHIV with the highest health needs. Moreover, this cluster was located in an urban and peri-urban area with high population density, where over 50% of the population in the community resided. These findings suggest the urgent need for an integrated approach that addresses multimorbidity in delivering care to this vulnerable community. Our study highlights an opportunity to leverage the existing HIV service infrastructure developed over the past few decades to address the needs of other chronic health conditions in these communities and to target their specific health needs.^33,34^

The distribution of categories of unmet health needs for PLHIV had similar spatial distributions, such that PLHIV engaged in care but sub-optimally treated (Score 2), those not engaged in care (Score 3) and those undiagnosed (Score 4) were spatially overlapping in the southern part of the study surveillance area. Distributions of health needs for people with hypertension and diabetes showed unique geospatial distributions. Individuals with sub-optimally treated or not engaged in care for hypertension were mainly distributed in the centre and norther parts of the surveillance area, whereas hypertensive individuals with the highest needs (Score 4) were clustered in the southern part of the surveillance area. Similarly, individuals with sub-optimally treated or not engaged in care for diabetes were mostly concentrated in the eastern and norther part of the surveillance area, whereas diabetic individuals with undiagnosed disease and not engaged in care showed similar distribution to the undiagnosed HIV and hypertensive patients and clustered in the southern part of the surveillance area.

Preliminary geospatial results from the Vukuzazi study using disease prevalence estimates found no spatial overlaps among the three chronic health conditions assessed in this study, HIV, hypertension, and diabetes.^6^ Similarly, results at national level suggested a lack of geographical overlap among disease prevalence estimations of HIV and other NCDs including cardiovascular diseases and diabetes in SA.^13^ These findings can be the result of disparities in the epidemic stages of these diseases disproportionately affecting different communities with distinct socioeconomic and demographic profiles. However, the implementation of a more granular epidemiological measures, like disease stages and the health needs emerging from these states, can uncover spatial associations and convergences of diseases affecting the same communities. Not all communities are experiencing the same health needs, and the identification of these uneven epidemiological landscape could support the design and implementation of prevention and control interventions targeting the specific health needs emerging from the affected community.

In this study, we identified different patterns of distribution for the various health needs for HIV, diabetes, and hypertension. We observed that consistent spatial patterns for all health needs were only occurring for HIV. PLHIV with sub-optimal treatment, not engaged in care, or undiagnosed, clustered in the southern part of the surveillance area, a location that is also experiencing high burden of the HIV epidemic with most of the HIV prevalence and incidence been concentrated in this area.^35^ Aligned with the 95-95-95 UNAIDS targets,^36^ these results highlight the need for the intensification of all 95’s targets in the identified area, with an increase in testing to identify those undiagnosed HIV-positive individuals, expansion of ART coverage to include those underserved individuals not engaged in care, and to sustain ART services to improve outcomes in individuals sub-optimally treated. However, it is important to note that PLHIV had the highest percentage of the population with the needs met in this HIV hyperendemic community,^17^{Singh, 2023 #42} with more than 78% of the HIV-positive participants having their health needs met (diagnosed, engaged in care, and optimally treated).

This result is consistent with the healthcare spending allocations of the country that are prioritizing the HIV epidemic and remains directed towards HIV interventions like ART.^37-39^ Conversely, there are limited finances available for other chronic health conditions.^39-41^ Consequently, results from the Vukuzazi study found a much lower percentage of the population with their health needs met for the NCDs included in the study, with only 6.9% of participants with diabetes and 41.8% of participants with hypertension having their health needs fully met.

Likewise, we identified clustered areas where about 15% of the total population had diabetes, and 17% had hypertension, with a disease sub-optimally treated or are participants not engaged in care, compared to the only 5% for PLHIV.

## Limitations

There are several limitations worth noting. Although a high rate of enrolment for a population-based multi-disease study was reported,^42^ non-systematic non-response in the Vukuzazi study including lower rates of enrolment by men could have biased the estimation of the health needs and their associations in directions hard to anticipate based on known differences between the sampled and unsampled population.^43^ In the Vukuzazi study elevated blood pressure and blood glucose were based on measurements conducted on a single day. Therefore, we acknowledge that people who screened positive for diabetes and hypertension required confirmatory testing prior to confirmation of diagnosis, and that this testing could rule out disease requiring immediate treatment. Thus, we may have overestimated the burden of undiagnosed disease.^44^ Furthermore, only three chronic disease conditions were considered in this study, and it must be taken into consideration that there are several other additional NCDs which have not been included in the Vukuzazi study including cancer, chronic respiratory diseases, mental health, and other NCDs, limiting our ability to draw comprehensive conclusions about population health and multimorbidity. Finally, the data analysed in this study are cross-sectional and so do not allow causal inference on their impact on morbidity and mortality.

## Conclusions

In this study, we identified distinct geospatial patterns in the distribution of health needs for HIV, diabetes, and hypertension in a HIV hyperendemic South African community. The identification of these patterns is essential for enhancing healthcare services tailored to the specific needs of these communities. In particular, there is a need to improve access to adequate care for individuals already diagnosed with these chronic diseases but lacking appropriate care. Notably, we found that individuals with the highest health needs for all three diseases were concentrated in the southern part of the surveillance area, indicating the potential benefits of integrating chronic disease management models developed for HIV care into the management of other chronic health conditions.^38^ Such integration could be a pivotal step towards improving healthcare delivery for vulnerable communities experiencing overlapping health conditions.

To achieve this goal, resource allocation should be based on geotargeting and tailored to the specific epidemiological needs of the affected community. Our findings underscore the importance of strategic resource allocation and efficient use of resources to improve health outcomes for populations with complex health needs. Therefore, our study provides valuable insights into the potential benefits of developing geographically targeted interventions aimed at improving access to health services and reducing the burden of overlapping health conditions.

## Data Availability

All data produced are available online at https://data.ahri.org/index.php/home

## ARTICLE INFORMATION

### Contributors

Concept and design: All authors.

Acquisition, analysis, or interpretation of data: All authors.

Drafting of the manuscript: DFC.

Critical revision of the manuscript for important intellectual content: All authors.

Statistical analysis: DFC, CD, US, SO.

Access to data and verified the data: DFC, US, SO.

### Data sharing statement

All data are available in public repositories: https://data.ahri.org/index.php/home

### Declaration of interest

The authors have no conflicts of interest to declare.

## Notes

### Competing Interest Statement

The authors have declared no competing interest.

